# Maternal Factors Promoting Normal Linear Growth of Children from Impoverished Rwandan Households

**DOI:** 10.1101/2024.03.11.24304145

**Authors:** Jean de Dieu Habimana, Noel Korukire, Jewett Sara, Eric Matsiko, Maryse Umugwaneza, Lawrence Rugema, Cyprien Munyanshongore

**Author notes:** **Corresponding author:** Jean de Dieu Habimana, School of Public Health, College of Medicine and Health Sciences, University of Rwanda, P.O. Box 3286 Kigali, Rwanda,.

## Abstract

**Background:** Linear growth faltering continues to be a pervasive public health concern, considering its persistent prevalence and repercussions. This study aimed to investigate potential maternal factors promoting normal linear growth among children from impoverished Rwandan households.

**Methods:** A structured questionnaire was used to collect data from 807 mother-child dyads selected using a multi-stage cluster sampling procedure. The main outcome variable was height-for-age Z-scores. The main predictors were the mother has income-generating activity, maternal education, maternal depression, household decision-making (HHDM), number of ANC visits, use of family planning method, family planning types, and mode of delivery. The potential confounding variables were child age, both parents working, good handwashing practice, owning a vegetable garden, and the total number of livestock. Univariate analysis was used to establish means, frequencies, and percentages; the Kruskal-Wallis, Mann-Whitney U, and Spearman Rank Correlation tests were used for bivariate analysis and robust linear regression for multivariable analysis.

**Results:** Maternal factors promoting normal linear growth of children were the presence of the mother’s income-generating activity (ꞵ=0.640 [0.0269−1.253], p-value=0.041), mother’s involvement in household decision-making (ꞵ=0.147 [0.080− 0.214], p-value<0.001), and higher frequency of ANC consultations (ꞵ=0.189 [0.025− 0.354], p-value=0.024). Additionally, a combination of household decision-making with ANC visit numbers predicted an increase in child linear growth (ꞵ=0.032 [0.019− 0.045], p-value<0.001).

**Conclusion:** Maternal factors such as maternal income-generating activity, maternal involvement in household decision-making, and increased number of ANC visits were found to promote normal child linear growth and can provide valuable information for shaping interventions and policies aimed at promoting child growth in the Rwandan community.

## Background

Linear growth faltering continues to be a pervasive public health issue that affects many children worldwide. In 2022, linear growth faltering affected approximately 148.1 million children globally, which amounts to 22.3% of all children under five years of age [1]. Sub-Saharan Africa was among the most affected, with 31.3%, thereby constituting a noteworthy concern [2]. Rwanda is among the countries with an alarming rate of linear growth faltering, with approximately 33.1% of children under five years affected in 2020 [3]. Thus, linear growth faltering continues to be a substantial health issue that requires further attention in Rwanda.

Linear growth faltering is the finest indicator of the accumulated effect of long-term undernutrition during the 1000 days of life [4]. It is explained as height-for-age Z-scores that fall below minus two standard deviations of the median height of World Health Organisation growth standards [5]. Linear growth faltering is not only an indicator of poor nutrition but also has short and long-term consequences such as reduced physical development, cognitive impairment and elevated proneness to infections [4], cardiovascular diseases [6], poor academic achievement, reduced physical resilience, and diminished economic productivity [7].

The negative outcomes of linear growth faltering can be notably devastating and cause irreversible damage, given that the first 1,000 days of life are a critical period of the child’s development [8]. In addition to that, linear growth faltering can also contribute to social and economic inequalities, perpetuating a cycle of poverty and malnutrition [9]. Short stature among mothers also increases the risks of having stunted children and spreads the vicious cycle of intergenerational linear growth faltering [10].

Effective strategies to promote normal linear growth include promoting maternal and child care, encouraging optimal feeding practices, providing early childhood education and stimulation, empowering women, promoting maternal psychosocial factors, and ensuring access to safe water and improved sanitation [11]. Addressing linear growth faltering also requires a multisectoral approach that includes collaboration of different sectors, including health, education, water and sanitation, and social protection [12]. With coordinated efforts, it is possible to promote normal linear growth and improve the overall health of millions of children worldwide.

The initiatives taken by Rwanda’s government and its stakeholders to contribute to the promotion of linear growth are evident. The prevalence of linear growth faltering among children has decreased from 44.2% in 2010 [13] to 33.1 in 2020 [3]. However, this decrease is still far from achieving the target of at least 19% by 2024, as proposed by the fourth health sector strategic plan for Rwanda [14]. This slight decrease testifies to the possibility of normal linear growth in some families. Thus, there is a need to explore other factors that could be associated with normal linear growth. The aim of this study was, therefore, to investigate potential maternal factors contributing to normal linear growth among children from impoverished Rwandan households.

## Methods

### Study setting, design, and population

This study was carried out in the districts of Kayonza, Nyaruguru, Burera, Rutsiro, and Gasabo from the provinces of eastern, southern, northern, western, and the City of Kigali, respectively. These districts were purposively selected due to their elevated prevalence of stunting.

This study used a cross-sectional design and a quantitative approach. We included children aged six to twenty-three months and their mothers who lived in the study districts. Inclusion criteria considered the child coming from a low-income family (category 1&2 of Ubudehe) [15], being a singleton, and being born full-term. Eligible participants who appeared to be too sick to participate were not considered to take part in the study.

The sample size of 807 participants was calculated using Fischer formula 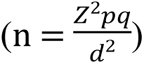 [16], considering a design effect of 2, a 20% loss to follow-up, and a precision level of 5%. In this formula, n indicates the sample size, and Z represents the abscissa of the normal distribution at a confidence level of 95% (Z=1.96), with the prevalence p of (33.1%), the complement of p, q = (1-p), and the desired accuracy of 5%.

We used a three-stage cluster sampling technique, where administrative villages served as the primary sampling unit, and households served as the unit of analysis. Extensive lists of all eligible candidates (mothers and children ages 6-23) who met the inclusion criteria were created from each chosen village in partnership with community health workers. After lists became available, systematic random sampling was used to select 807 mother-child dyads.

Qualified and experienced enumerators were recruited and trained for three days: one day for theory, one day for practice, comprising the pretest, and one day reserved for the adjustment of the data collection tool. Community health workers from the visited villages helped us to identify eligible households. Twenty homes in one sector of the Gasabo district participated in a pretest; the results were used to update and correct the data collection instruments. We chose one sector within the Gasabo district due to the same attributes as the research area. However, the study did not cover this sector.

Data was collected from 30^th^ August to 23^rd^ September 2021 by twenty-five enumerators grouped into five teams. Each team was composed of six enumerators and one supervisor. Enumerators worked in pairs to help each other during the data collection. One team composed of three pairs was assigned to collect data in one District and coordinated with one supervisor. Data collection was conducted in the participants’ households. In order to avoid data entry errors and save time for data entry, quantitative data were gathered using a data collection tool installed in tablets with an electronic data management system called Kobo Collect.

A portable stadiometer was used to measure the child’s length. The remaining data were gathered through a structured questionnaire. All day-to-day collected data were sent to the central system, where quality checks were done daily to ensure completeness of data. The primary investigator and the supervisors oversaw every aspect of the data-gathering process.

### Study variables

The main outcome variable of this study was linear growth, and it was presented in the form of height-for-age Z-scores as a continuous variable. The main predictors of this study were the mother having income-generating activity, maternal education, maternal depression, household decision-making (HHDM), number of ANC visits, use of family planning method, family planning types, and mode of delivery. The potential confounding variables were child age, engagement in works for both parents, good handwashing practice, lack of vegetable gardens, and the total number of livestock (inclusive of chicken, goats, and cattle).

To assess maternal depression, we used the EPDS, a ten-item tool with a 0-3 Likert scale and a maximum score of 30 [17]. When it comes to interpretation, the higher the value, the worse the depressive symptoms. The EPDS tool has been widely used throughout sub-Saharan Africa [18]. In addition, it has been validated for examining the extent and potential risks of depression during the perinatal period in numerous countries and languages [18]. It has also been verified as an antenatal depression screening tool in a previous Rwandan study [19]. In the present study, the EPDS was shown to be reliable, with Cronbach’s alpha value was 0.840.

For household decision-making, we used a three-item scale to measure the women’s household decision-making, with the total score ranging between zero and nine [20]. For interpretation, a higher score reflects more maternal involvement in decision-making. For this study, the tool was deemed reliable, attaining a Cronbach’s Alpha value of 0.895.

### Data analysis

Data were analyzed using STATA 15 software. In order to export the data into stata for final cleaning and analysis, a data dictionary was developed. After data cleaning, the Shapiro-Wilk Test was used to check for the normality of the dependent variable. Initially, descriptive analysis helped to summarize study variables using frequencies and percentages or means and standard deviations.

The next level involved specifying significant variables considering the main outcome variable. Given that the outcome variable was not normally distributed, we opted to use the Kruskal-Wallis, Mann-Whitney U, and Spearman Rank Correlation tests for bivariate analysis and Huber’s approach for robust linear regression for multivariable analysis. This approach allows for vigorous estimation of the model coefficients, accommodating potential outliers and providing reliable findings in the presence of diverse variable types [21]. Only significant variables from bivariate analysis (p-value ≤ 0.10) were included in the robust regression analysis.

The robust regression analysis models were fitted in three steps. In the first step (Model I), significant variables with p-value ≤0.10 from the bivariate analysis were entered one by one. In the second step (Model II), the significant variables (p-value ≤ 0.05) related to maternal factors were entered one by one into the Model after controlling for other significant factors from Model I, including child age, good and washing practice, and the number of livestock. In the third stage (Model III), we entered all significant variables from Model II with p≤0.05 after controlling the same variables as in Model II. Only significant variables were considered independent factors of normal linear growth. The results were reported as a beta coefficient (ꞵ) with a 95% confidence interval (CI) and p-value ≤ 0.05. In addition, after finding independently significant variables, we thoroughly analyzed their combined effects. To be more precise, we created two separate models to look into the relationship between a combination of household decision-making and ANC visit numbers, a combination of household decision-making and the mother having income-generating activity, a combination of the numbers of ANC visits and the mother having income-generating activity, as well as the interaction of all three variables together. The process used to construct these interaction models followed the same exacting guidelines as our earlier analysis.

### Ethical considerations

The Ethical approval was issued by the Institutional Review Board of the College of Medicine and Health Sciences, University of Rwanda (N°. 335/CMHS IRB 2021, which amended N°. 178/CMHS IRB 2022). Additionally, a research permit was obtained from the Rwanda Ministry of Local Governance and the National Institute of Statistics of Rwanda. In addition, the mothers signed their written consent to participate voluntarily.

## Results

### Linear growth among children

Height-for-age Z-scores, reflecting linear growth for the sample, exhibit a mean of -1.38 ±1.78. As shown in Figure 1, curve A represents a normal distribution, while curve B reflects the study sample’s growth curve. Besides, curve B demonstrates a left-skewed tendency with a concentration of children aged 6-23 months from impoverished Rwandan households reflecting relatively lower height-for-age Z-scores. This negative value suggests a tendency towards linear growth faltering. However, even though the present mean displays a negative skew, a large number of children still fell within the normal range (above minus two standard deviations), indicating that a significant number of children had a likelihood of improving their height-for-age Z-scores despite the unhurried improvement.

**Figure 1.**
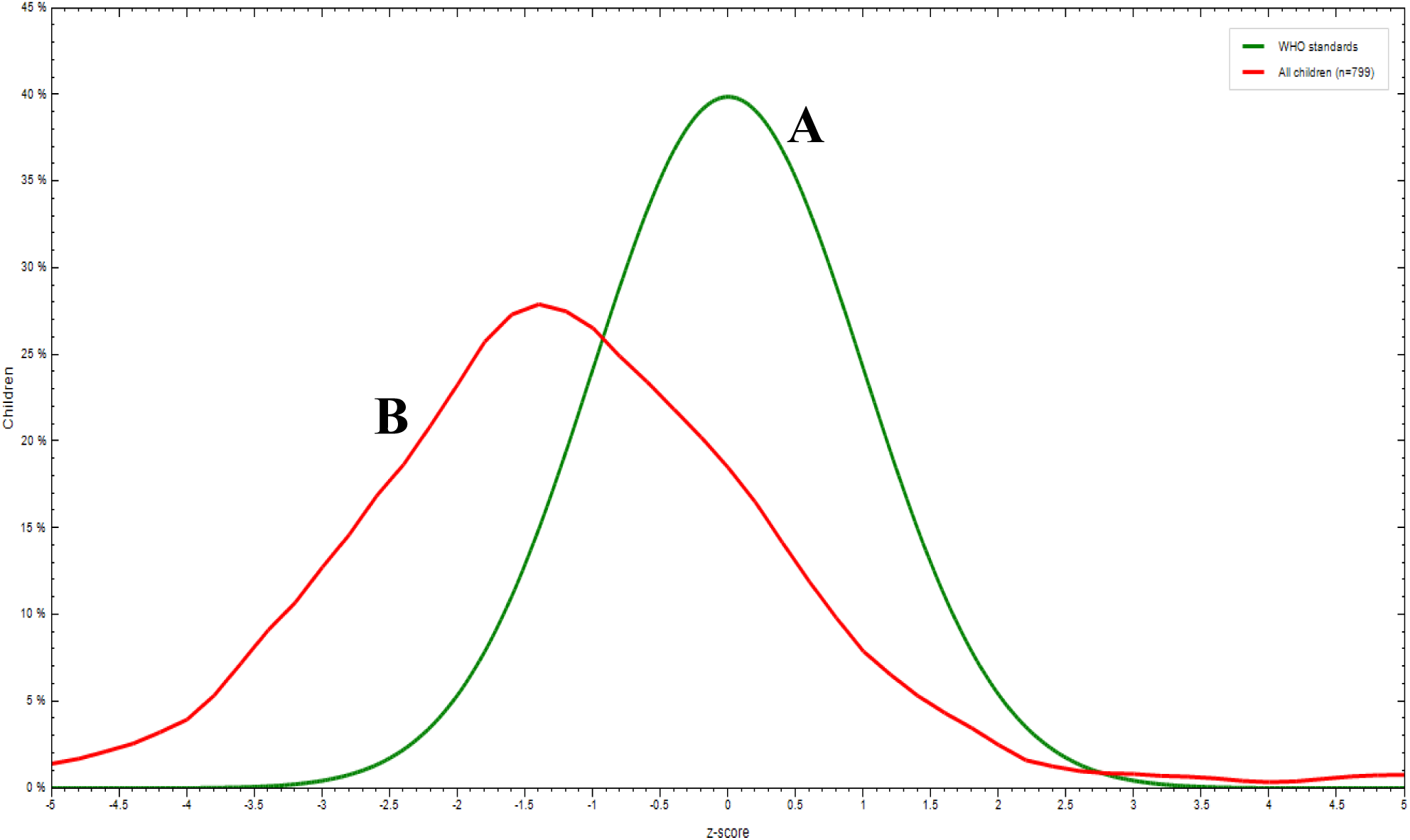
Height-for-age Z-scores (linear growth)

### Sociodemographic characteristics of study participants

As detailed in Table 1, of the mothers who completed the survey, the mean EPDS score was 10.84 ±5.91 out of a maximum of 30. This mean demonstrates a moderate level of depressive syndromes within the study population. Furthermore, the mean HHDM score was 7.44 ±2.09 out of a maximum of 10. The mean here shows a relatively high level of involvement of the mothers in household decision-making among the study population.

**Table 1:** Sociodemographic characteristics of study participants (N=807).

| Variables | Mean± SD | Frequency | Per cent | Bivariate analysis |  |
| --- | --- | --- | --- | --- | --- |
|  |  |  |  | ρ/ U/H | p-value |
| Child age | 14.171±5.13 | - | - | -0.193 | **<0.001** |
| EPDS | 10.84 ±5.91 | - | - | -0.101 | **0.004** |
| Household decision-making scores | 7.44 ±2.089 | - | - | 0.217 | **<0.001** |
| ANC visits numbers | 3.51 ±0.855 | - | - | 0.113 | **0.001** |
| Maternal education |  |  |  |  |  |
| No formal education | - | 89 | 11.03 | 10.714 | **0.005** |
| Primary | - | 505 | 62.58 |  |  |
| Post-primary | - | 213 | 26.39 |  |  |
| Mother has income-generating activities. |  |  |  |  |  |
| No | - | 745 | 92.3 | -2.008 | **0.045** |
| Yes | - | 62 | 7.7 |  |  |
| Mother works alone |  |  |  |  |  |
| No | - | 30 | 96.3 | -2.351 | **0.019** |
| Yes | - | 777 | 3.7 |  |  |
| Use of family planning method |  |  |  |  |  |
| No | - | 401 | 49.7 | 0.708 | 0.479 |
| Yes | - | 406 | 50.3 |  |  |
| Breast discomfort during lactation |  |  |  |  |  |
| No | - | 696 | 86.2 | 2.085 | **0.037** |
| Yes | - | 111 | 13.8 |  |  |
| Family planning types |  |  |  |  |  |
| No family planning before | - | 402 | 49.8 | 1.508 | 0.470 |
| Modern | - | 381 | 47.2 |  |  |
| Traditional | - | 24 | 3.0 |  |  |
| Delivery mode |  |  |  |  |  |
| Not assisted by health facility | - | 15 | 1.9 | 11.24 | **0.004** |
| Vaginal delivery from a health | - | 703 | 87.1 |  |  |
| Cesarean | - | 89 | 11.0 |  |  |
| Total number of livestock | 1.48 ±2.414 | - | - | 0.080 | **0.024** |
| Vegetable garden possession |  |  |  |  |  |
| No | - | 395 | 48.9 | 1.572 | 0.116 |
| Yes | - | 412 | 51.1 |  |  |
| Good handwashing practice |  |  |  |  |  |
| No | - | 160 | 25.9 | -6.097 | **<0.001** |
| Yes | - | 458 | 74.1 |  |  |
| Both parent working |  |  |  |  |  |
| No | - | 111 | 13.7 | 0.175 | 0.861 |
| Yes | - | 696 | 86.3 |  |  |

\* **ρ** is the coefficient for Spearman rank correlation, **U** for the Mann-Whitney U test, and **H** for the Kruskal-Wallis test.

In relation to socioeconomic status, the mean livestock owned by households was 1.48 domestic animals. Only 7.7% of mothers said they had income-generating activities, with even fewer 3.7%) claiming to be only the ones who work in the household, and 11.03% did not receive formal education.

In terms of reproductive health, around half of all women (49.7%) did not use contraceptive methods before getting pregnant. The mean number of ANC visits was 3.50 ±0.863, while Rwanda recommends at least 4 ANC visits. Further, most mothers gave birth in a health facility, with only 1.9% reporting giving birth outside a health facility. Post-delivery, 13.8% of mothers claimed to experience breast discomfort during lactation. In addition, about half of all surveyed households (51.1%) had a vegetable garden, while 74.1% demonstrated good handwashing practices, and 86.3% reported having both parents working.

Using the same table, we examined the relationship between the sociodemographic characteristics of mothers and the normal linear growth of children. The following variables were significant (p-value < 0.05): child age (p-value<0.001), maternal education (p-value= 0.005), EPDS (p-value=0.004) and HHDM (p-value<0.001), ANC visits numbers (p-value=0.001), the mother has income-generating activities (p-value<0.001), the mother works alone (p-value= 0.019), experience breast discomfort during lactation (p-value= 0.037), mode of delivery (p-value= 0.004), the total number of livestock (p-value=0.024), and good handwashing practice(p-value<0.001).

### Multivariable analysis: maternal factors promoting a normal linear growth of children

Table 2 shows the results of multivariable analysis into three models, with the final model results presented here. The results suggest that maternal engagement in income generation, higher scores of maternal involvement in household decision-making, and more ANC visits are all positively associated with higher height-for-age Z-scores. To be more specific, the mother’s being engaged in income-generating activities increased by 0.640 to the Z-scores of height-for-age of the child (ꞵ=0.640 [0.0269−1.253], p-value= 0.041). In addition, with an increase in one unit of household decision-making, there is an increase of 0.147 in Z-scores of height-for-age of the child (ꞵ=0.147 [0.080− 0.214], p-value< 0.001). Further, in line with the increase in one antenatal care visit for the woman, there is an additional 0.189 units to the Z-score of height-for-age (ꞵ=0.189 [0.025− 0.354], p-value= 0.024).

**Table 2:**
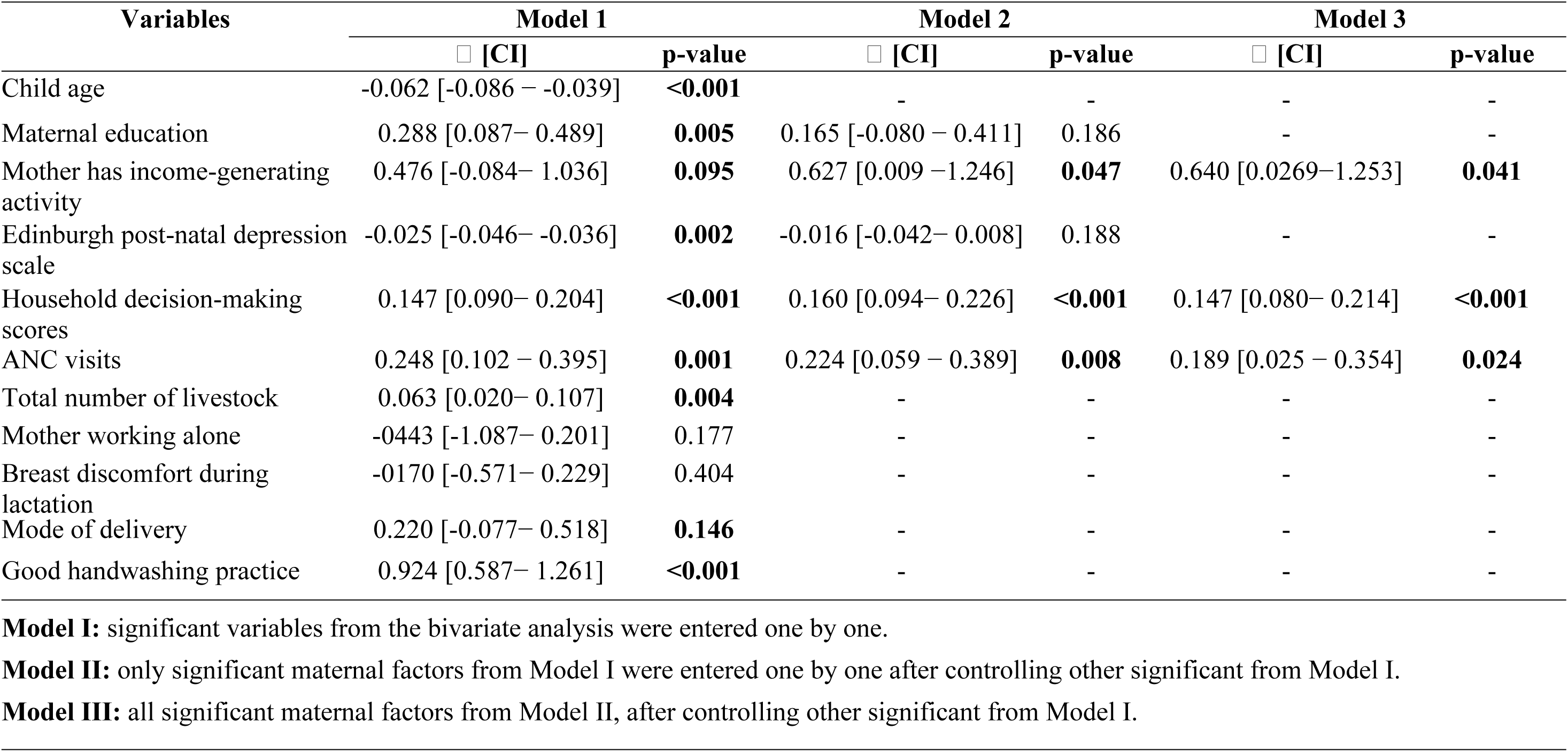
Multivariable analysis: factors promoting normal linear growth of children (N=807).

Table 3 presents the combined effects of all significant variables. The findings showed that a combination of HHDM and ANC visits was significant (ꞵ=0.032 [0.019− 0.045], p-value<0.001). This coefficient means that for each unit increase in the combination of household decision-making scores and number of ANC visits, a corresponding increase of 0.032 units in linear growth was anticipated.

**Table 3:** Combined effect between significant variables that promote linear growth (N=807).

| Variables | Model 1 |  | Model 2 |  | Model 3 |  |
| --- | --- | --- | --- | --- | --- | --- |
|  | □ [CI] | p-value | □ [CI] | p-value | □ [CI] | p-value |
| Household decision-making and number of ANC visits | 0.031 [0.020– 0.042] | **<0.001** | 0.031 [**0.018– 0.044**] | **<0.001** | 0.032 [0.019– 0.045] | **<0.001** |
| Household decision-making and the mother has income-generating activity | 0.071 [0.001– 0.1419] | **0.048** | 0.084 [0.004– 0.164] | **0.039** | 0.117 [-0.203– 0.438] | 0.473 |
| Number of ANC visits, and the mother has income-generating activity | 0.140 [-0.023 – 0.304] | **0.093** | 0.206 [0.017– 0.396] | **0.033** | 0.410 [-0.567 – 1.387] | 0.410 |
| Household decision-making, number of ANC visits, and the mother has income-generating | 0.019 [-0.001– 0.039] | **0.056** | 0.025 [0.001 – 0.049] | **0.043** | -0.060[-0.219 –0.098] | 0.456 |

**Model I:** combined significant variables from multivariable analysis were entered one by one.
**Model II:** combined significant factors from Model I were entered one by one after controlling the same factors.
**Model III:** all significant maternal factors from Model II were entered one by one after controlling the same factors.

## Discussion

Linear growth as expressed by height-for-age Z-scores for children in this study was, on average-1.38 ± 1.78. Compared to the expected average for their age group, this mean is relatively lower. The fact that the study sample was, on average, shorter than the reference population may raise concerns about the trend of linear growth faltering among children from poor Rwandan households. This trend is supported by numerous studies that have been conducted previously [22,23]. On a more optimistic note, despite this negative tendency, there was still a large number of children that fell in the normal range of linear growth since the mean did not fall under minus two. The present mean aligns with a study that indicates the possibility of normal linear growth despite belonging to low socioeconomic families [24].

The present study explored maternal factors that could promote child linear growth. The mean of 10.84 ± 5.91 for the EPDS score indicates an elevated level of depressive syndromes within the studied population. These findings are consistent with other studies conducted in Rwanda, which showed a high prevalence of antenatal depressive symptoms [18,25]. Despite the elevated prevalence of maternal depressive symptoms, we did not find any statistically significant associations with childhood linear growth.

Unlike maternal depression, this study did find a positive association between household decision-making and linear growth. The mean HHDM) scores of 7.44 ± 2.089 suggest that mothers in the study population felt relatively empowered in relation to household decision-making. This empowerment implies that an empowered decision-making process within households may contribute positively to the linear growth of children[26]. Our study findings were consistent with studies conducted in Ethiopia [27,28] and Nigeria [29], where poor maternal implication in household decision-making decreased height-for-age Z-scores for children.

The results of this study revealed a substantial positive correlation between the number of ANC visits for women and the linear growth of the children. This association could be justified by the fact that the parents who regularly attended ANC received health and nutrition education. In addition, ANC was a window for them to be enrolled in other programs set by the government that target improving their lives. Unfortunately, this was not explored. However, another study conducted in Sub-Saharan Africa has also found this association [24] and points to the potential role of maternal healthcare utilization during pregnancy in promoting child growth.

Mothers being engaged in income-generating activities increased the height-for-age Z-scores of the child in this study. This relationship may be due to improved access to socioeconomic resources that contribute to the optimization of purchase power, better food, healthcare, and living conditions. Another study conducted in Cambodia found that increased income can improve a child’s access to healthcare and better nutrition [30]. Another study from Bangladesh established a positive relationship between maternal employment and the overall economic well-being of the family with long-term favourable benefits on the child’s development [31]. That being said, some studies have found the opposite when a mother is involved in any activity that can generate income [32,33]. The reason behind this could be the reduced time to allocate to children and hence reduced child care [34].

From this analysis, we explored the combined effects of both household decision-making scores and the number of antenatal care (ANC) visits on linear growth. This positive association suggests that the effect of maternal involvement in household decision-making on child linear growth is more pronounced in the presence of higher levels of ANC visit numbers. The findings from this study imply that the joint influence of household decision-making scores and ANC visit numbers significantly strengthens their combined effect on child linear growth. In essence, the positive impact of household decision-making scores on linear growth is more pronounced when there are higher levels of ANC consultations.

This study has some limitations. First, the cross-sectional nature limits our ability to determine the causal-effect inference. Secondly, apart from anthropometric measurements, many of the variables evaluated were self-reported, which may have been subject to social desirability bias. Third, we did not measure other factors, such as nutrition biomarkers, that can affect linear growth as well.

In conclusion, the results from this study collectively elaborate the multifaceted nature of factors promoting child linear growth, including maternal implication in household decision-making, increased number of ANC consultations, and mother’s income-generating. These results contribute valuable information for shaping interventions and policies aimed at improving child nutrition and growth in the studied community.

## Data Availability

The data will be accessible to anyone who desires to access them for scientific reasons. The request should be made through the corresponding author.

